# IMPACT OF THE CORONAVIRUS DISEASE 2019 PANDEMIC ON SURGICAL VOLUME IN JAPAN: A COHORT STUDY USING ADMINISTRATIVE DATA

**DOI:** 10.1101/2020.11.18.20233882

**Authors:** Takuya Okuno, Daisuke Takada, Shin Jung-ho, Tetsuji Morishita, Hisashi Itoshima, Susumu Kunisawa, Yuichi Imanaka

## Abstract

**Background:** Internationally, the Coronavirus Disease (COVID-19) pandemic has caused unprecedented challenges for surgical staff to minimise the exposure to COVID-19 or save medical resources without harmful outcomes for patients, in accordance with the statement of each surgical society. However, no research has empirically validated declines in Japanese surgical volume or compared decrease rates of surgeries during the COVID-19 pandemic.

**Material and Methods:** We extracted 672,772 available cases of patients aged > 15 years who were discharged between July 1, 2018, and June 30, 2020. After categorisation of surgery, we calculated descriptive statistics to compare the year-over-year trend and conducted interrupted time series analysis to validate the decline.

**Results:** The year-over-year trend of all eight surgical categories decreased from April 2020 and reached a minimum in May 2020 (May: abdominal, 68.4%; thoracic, 85.8%; genitourinary, 78.6%; cardiovascular, 90.8%; neurosurgical, 69.1%; orthopaedic, 62.4%; ophthalmologic, 52.0%; ear/nose/throat, 27.3%). Interrupted time series analysis showed no significant trends in oncological and critical benign surgeries.

**Conclusion:** We demonstrated and validated a trend of reduction in surgical volume in Japan using administrative data applying interrupted time series analyses. Low priority surgeries, as categorised by the statement of each society, showed obvious and statistically significant declines in case volume during the COVID-19 pandemic.

## 1. Introduction

The rapid spread of the coronavirus disease (COVID-19), which first appeared in Wuhan, China, has been increasing and disrupting healthcare systems worldwide.^1^ During the COVID-19 pandemic, there have been over 4.5 million cases and over 300,000 deaths internationally, which has caused unprecedented challenges for surgical staff.^2-4^ To minimise exposure to COVID-19 and save medical resources to prevent harmful outcomes for patients, cancellation or triage of surgeries are recommended.^3-5^ Patients with critical disease and cancers should still receive surgery as usual, whereas cases with benign diseases can be postponed.^6-8^

Previous studies suggest that there are wide variations of trends in the affected number of surgical procedures depending on the country and time period.^3,4,9^ For example, estimates of cancelled or postponed surgeries during the first wave of the COVID-19 pandemic in Japan were about 1.4 million cases (73% of the total), similar to other high-income countries.^3^ Of these, approximately 98,000 were oncological surgeries with a cancellation rate of 30% and the remaining 1,253,000 cases were benign surgeries with a cancellation rate of 84%.^3^ Referring to this estimate, the Japanese Surgical Society (JSS, which consists of 10 major surgical societies in Japan) announced the importance of triage by the patients’ severity and surgical recovery planning for the backlog of surgeries.^10^ A comprehensive surgical and medical care strategy is also recommended to prevent potential adverse health implications caused by surgical delays.^11^

However, there are no studies on the trends of surgeries during the pandemic in Japan, one of the regions most affected by COVID-19 at the earliest stage of the pandemic and where the first wave of the pandemic converged most quickly (Figure 1). Several publications have dealt with the issue of triaging elective surgeries, but the types of surgeries affected remains unclear. In this report, we performed a retrospective cohort study discussing the impact of the first wave of the COVID-19 pandemic on Japanese elective surgical procedures by major surgical types using administrative data in Japan. We sought to identify which surgical areas were the most affected by the COVID-19, and to statistically evaluate the decline in major operative procedures, referring to the priorities announced by each surgical society.

**Figure 1.**
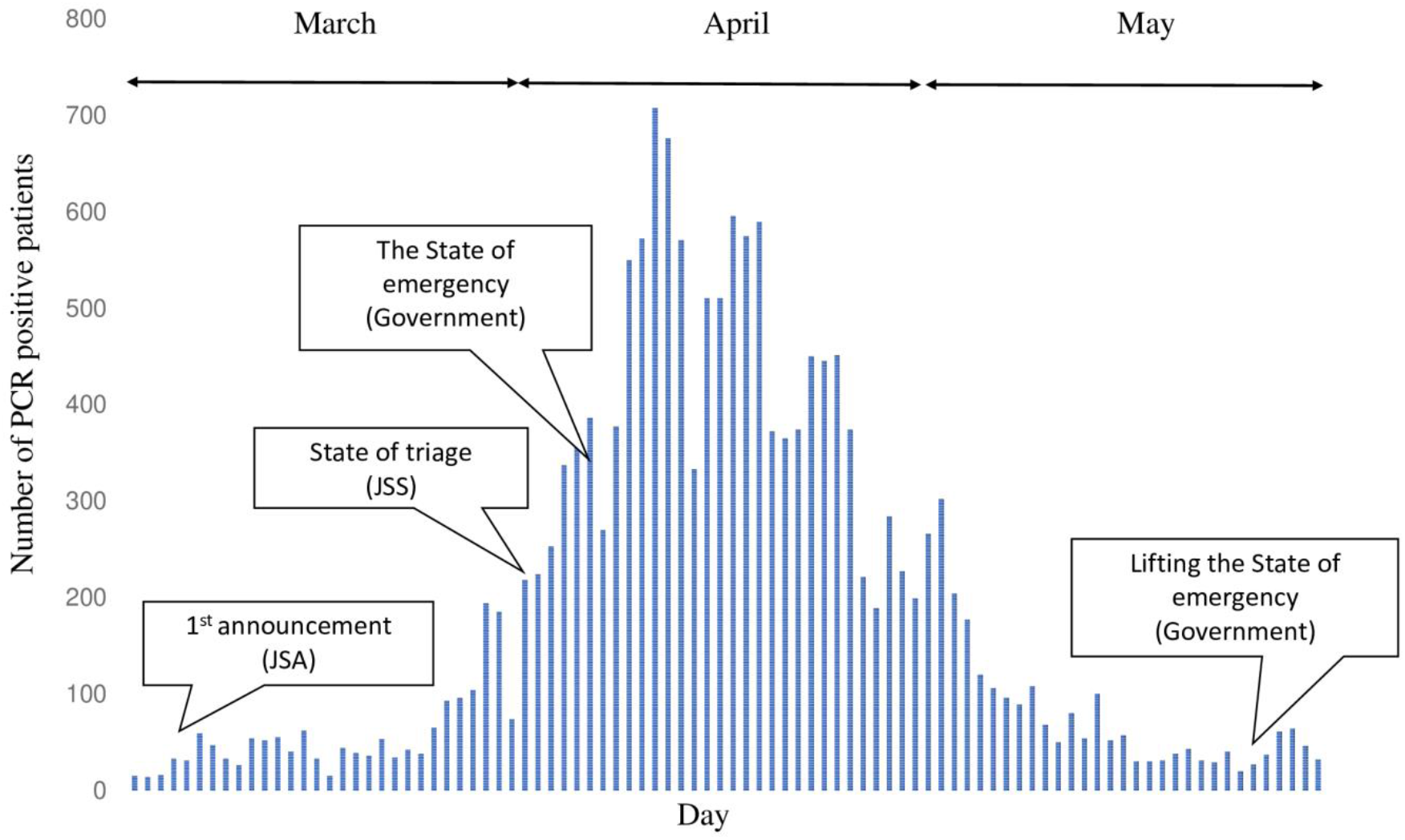
Trend of PCR tests for COVID-19 positive patients in Japan between March 1, 2020 and May 31, 2020, with major statement by the JSA, JSS, and Japanese government JSA, Japanese Society of Anaesthesiologists; JSS, Japan Surgical Society; PCR, Polymerase Chain Reaction

## 2. Methods

### 2.1. Data source

We used Diagnosis Procedure Combination (DPC) data from the Quality Indicator/Improvement Project (QIP) database, the programme administered by the Department of Healthcare Economics and Quality Management, Kyoto University. QIP participant hospitals voluntarily provide claim data, the DPC data, to improve their quality of care and system using quality indicators. There were over 200 QIP participant hospitals all over Japan including both public and private hospitals with various sizes: the number of general beds, which are hospital beds that are not psychiatric, infectious diseases, and tuberculosis beds, according to the Japanese classification of hospital beds, ranged from 30 to 1,151 in 2019.

The DPC/per-diem payment system (PDPS) is a Japanese prospective payment system applied to acute care hospitals, which is comparable to diagnosis-related databases in the United States.^12,13^ A total of 1,730 hospitals adopted the DPC/PDPS in 2018, which accounted for 54% of all general beds in Japanese hospitals.^14,15^ DPA data do not include information on laboratory findings and reasons for surgery; instead provide information included in the discharge summary, such as primary diagnosis, comorbidities (identified by using the International Classification of Diseases, 10th Revision [ICD-10] codes), type of surgery, drug or device prescriptions, and codes corresponding to the medical procedures performed.

### 2.2. Study population

Among QIP hospitals, we detected 254 hospitals, 672,772 cases of patients over 15 years old with surgery whose discharge summary was available from between July 1, 2018, and June 30, 2020, which was used for interrupted time series analysis (ITS) analysis. Figure 2 shows the flow chart of selection processes for each analysis. After identification of the population for ITS analysis, to see the trend of the number of surgical cases during the first wave of the pandemic, we compared patients who were admitted and discharged between July 1, 2018 and June 30, 2019 with those between July 1, 2019 and June 30, 2020.

**Figure 2.**
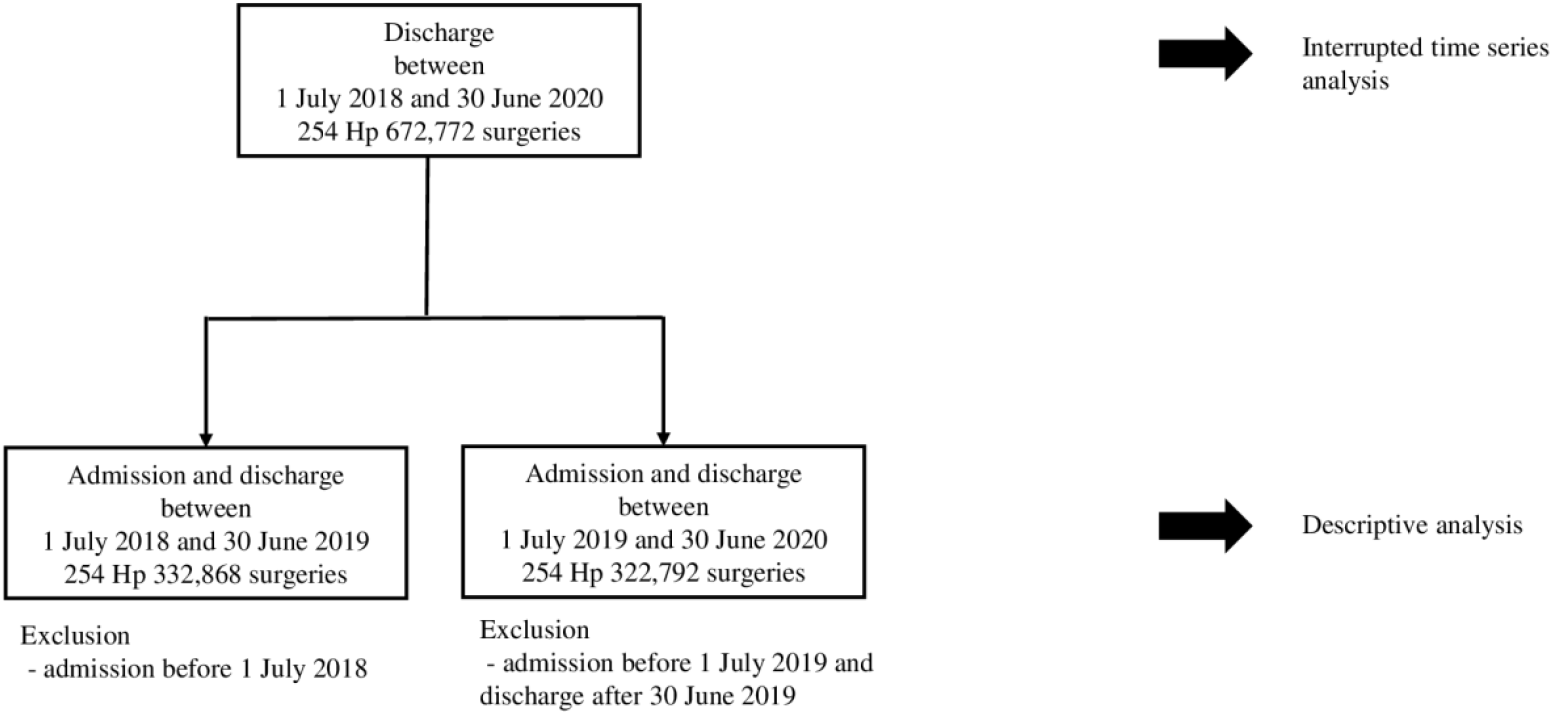
Flowchart depicting patient progression through the present study eligibility criteria HP, hospitals

### 2.3. Variables and surgical type

For patients’ background characteristics, we referred to their sex, age, and length of stay (LOS), as well as the Charlson and Elixhauser comorbidity indices.^16^ We also identified the type of surgery from the main surgery during admission. Categorisation was based on the major surgical areas and previous reports^5,9^: abdominal (stomach cancer surgery; colorectal cancer surgery; hepato-pancreatic-biliary [HPB], cancer surgery; benign popular including hernia surgery, cholecystectomy, and appendicectomy; other benign surgeries), thoracic (lung cancer, breast cancer, and benign surgeries), genitourinary (bladder cancer; prostate cancer; uterine/adnexa cancer; benign popular surgeries including hysterectomy, salpingo-oophorectomy; other benign surgeries; caesarean section), cardiovascular (valve surgery, aortic surgery, coronary artery bypass grafting [CABG], dialysis fistula, other), neurosurgery (intracranial surgery including hematoma, vascular, and tumour; spine surgery; other), orthopaedic (fracture-related surgery; arthroplasty; other), ophthalmology (popular surgery including cataract, vitreous, and blepharoplasty; other), and ear/nose/throat (ENT: popular surgery including tonsillectomy, endoscopic sinus surgery, and tympanoplasty; other). Due to the wide variety of surgical procedures, we created the category of popular surgery defined as the top three most common surgeries in Japan based on the National Database of Health Insurance Claims and Specific Health Checkups of Japan (NDB), which covers the medical claim information of the entire Japanese population of 127 million.^17^ Details of surgical classification are shown by K codes, which are the claims codes for surgery, in supplemental Table 1.

**Table 1.** Monthly cases by surgical type between July 2018 and June 2020 for interrupted time series analysis.

|  |  | Jul | Aug | Sep | Oct | Nov | Dec | Jan | Feb | Mar | Apr | May | Jun |
| --- | --- | --- | --- | --- | --- | --- | --- | --- | --- | --- | --- | --- | --- |
| Abdominal | July 2018 to June 2019 | 4997 | 5317 | 4761 | 5169 | 5162 | 5544 | 4056 | 5019 | 5449 | 5021 | 4550 | 5132 |
|  | July 2019 to June 2020 | 5233 | 5304 | 5010 | 5131 | 5088 | 5659 | 4123 | 5104 | 5236 | 4434 | 3495 | 4312 |
| Thoracic | July 2018 to June 2019 | 1717 | 1721 | 1651 | 1775 | 1796 | 1978 | 1381 | 1655 | 1777 | 1797 | 1462 | 1694 |
|  | July 2019 to June 2020 | 1678 | 1739 | 1605 | 1768 | 1765 | 2099 | 1389 | 1785 | 1783 | 1638 | 1330 | 1528 |
| Genitourinary | July 2018 to June 2019 | 6117 | 6571 | 5901 | 6334 | 6093 | 6415 | 5133 | 5924 | 6393 | 6136 | 5474 | 6284 |
|  | July 2019 to June 2020 | 6352 | 6635 | 5825 | 6233 | 6204 | 6864 | 5210 | 6124 | 6289 | 5678 | 4722 | 5356 |
| Cardiovascular | July 2018 to June 2019 | 2415 | 2292 | 2264 | 2259 | 2264 | 2419 | 1685 | 2047 | 2307 | 2258 | 1953 | 2304 |
|  | July 2019 to June 2020 | 2496 | 2291 | 2148 | 2227 | 2295 | 2531 | 1777 | 2247 | 2141 | 2059 | 1601 | 1981 |
| Neurosurgical | July 2018 to June 2019 | 2007 | 1952 | 1760 | 1797 | 1887 | 2107 | 1299 | 1728 | 2003 | 1793 | 1613 | 1900 |
|  | July 2019 to June 2020 | 2016 | 1996 | 1774 | 1810 | 1766 | 2159 | 1384 | 1813 | 1884 | 1729 | 1254 | 1611 |
| Orthopaedic | July 2018 to June 2019 | 4894 | 5427 | 4669 | 5003 | 5119 | 5578 | 4023 | 5128 | 5889 | 5076 | 4582 | 5036 |
|  | July 2019 to June 2020 | 5349 | 5697 | 4728 | 5228 | 5386 | 5934 | 4283 | 5511 | 5675 | 4324 | 3433 | 3905 |
| Ophthalmologic | July 2018 to June 2019 | 5462 | 5596 | 5007 | 5632 | 6227 | 5160 | 4866 | 5587 | 6007 | 5425 | 5405 | 6158 |
|  | July 2019 to June 2020 | 6153 | 5629 | 5095 | 5763 | 6490 | 5431 | 5408 | 5682 | 5734 | 4567 | 2978 | 4345 |
| Eye/Nose/Throat | July 2018 to June 2019 | 1039 | 1147 | 1015 | 1069 | 1099 | 1145 | 843 | 1104 | 1180 | 1094 | 947 | 1088 |
|  | July 2019 to June 2020 | 1168 | 1200 | 1154 | 1142 | 1167 | 1295 | 943 | 1139 | 1148 | 683 | 280 | 612 |

### 2.4. Statistics

#### 2.4.1. Descriptive statistics

The year-over-year trend of the case volume per month by the eight types of surgery between July 1, 2019 and June 30, 2020 compared to that between July 1, 2018 and June 30, 2019 is presented in Figure 3, using line graphs. Especially, details of each surgical category in March, April, and May, which appear to be the first wave COVID-19 pandemic, were calculated.

**Figure 3.**
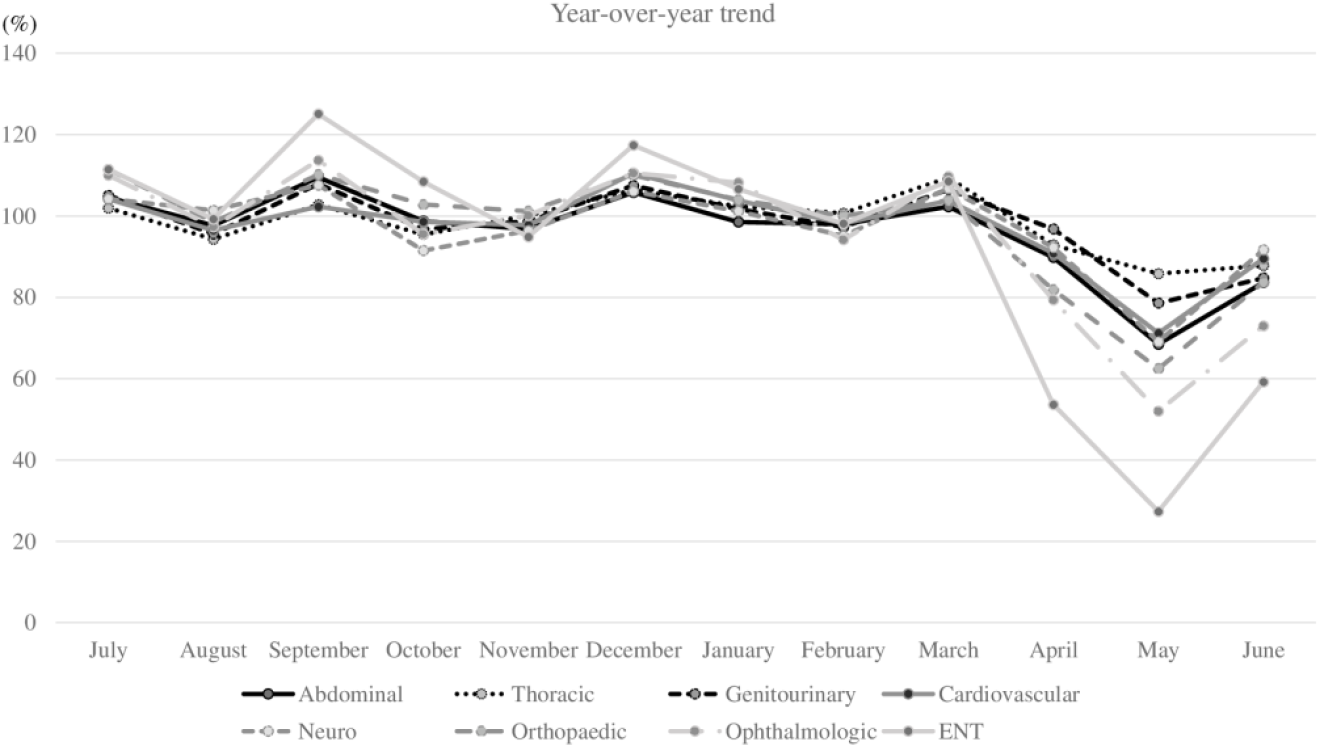
The year-over-year trend in the number of surgeries by the eight categories ENT, Ear/Nose/Throat

#### 2.4.2. Interrupted time series analyses for changes in case rates

We used ITS, including segmented regressions, to ascertain volume changes due to the COVID-19 pandemic on population-level admissions. We statistically tested the changes of the volume of surgeries based on the date of discharge adjusted for seasonality through a Fourier term.^18^ We regarded comparing the volume of scheduled admissions with surgeries per month using the date of discharge rather than admission, to be appropriate because DPC data were generated after patients’ discharge. We hypothesise that COVID-19 would have affected the level change of scheduled admission volume especially after April 2020 as an implemented point, in which the Japanese Society of Anaesthesiologists and the JSS requested surgeons to triage some elective surgeries. We divided our datasets into two periods for ITS analysis: before the JSS surgical-triage-statement (July 2018 to March 2020) and after that (April 2020 to June 2020). Monthly case volumes of scheduled admission with surgery are presented in the figures using line graphs. Baseline demographics for the population for ITS analysis are listed in Supplemental tables 2∼10. Age was expressed as mean ± SD and LOS as median [interquartile range] and were compared using Mann-Whitney U tests. Categorical variables were expressed as percentages and compared using χ^2^ tests. The level of statistical significance was set at p < 0.05 (two-tailed). Statistical analyses were performed with R version 4.0.2 (R Foundation for Statistical Computing, Vienna, Austria).

The study protocol was approved by the Ethics Committee of Kyoto University Graduate School, Kyoto, Japan (R0135). This study was conducted in accordance with the ethical guidelines for medical and health research involving human participants issued by the Japanese National Government. The data were anonymised, and the requirement for informed consent was waived.

## 3. Results

### 3.1. Overview of entire surgery

Figure 3 displays the year-over-year trend of the case volume per month of each surgical type based on the date of admission. Every type of surgical volume, especially ophthalmologic and ENT surgeries, tended to decrease from April and reached a minimum in May (May: abdominal, 68.4%; thoracic, 85.8%; genitourinary, 78.6%; cardiovascular, 90.8%; neurosurgical, 69.1%; orthopaedic, 62.4%; ophthalmologic, 52.0%; ENT, 27.3%; supplemental Table 2). Table 1 shows the monthly cases by the eight surgical categories based on the date of discharge, which was calculated for ITS analysis, and also indicated the smallest number of cases in May for all surgical types.

### 3.2. Abdominal surgery

Prior year comparison of stomach, colorectal, and HPB cancer were almost the same in March and April. However, benign surgeries were considerably lower in April and May than in the previous year (supplemental Figure 1: popular, 53.9%; other, 71.6%). According to the ITS analysis, the surgical volume of oncological surgeries was not statistically affected by COVID-19, whereas benign surgeries were affected (Figure 4: benign popular, p = 0.007; benign other, p = 0.018).

**Figure 4.**
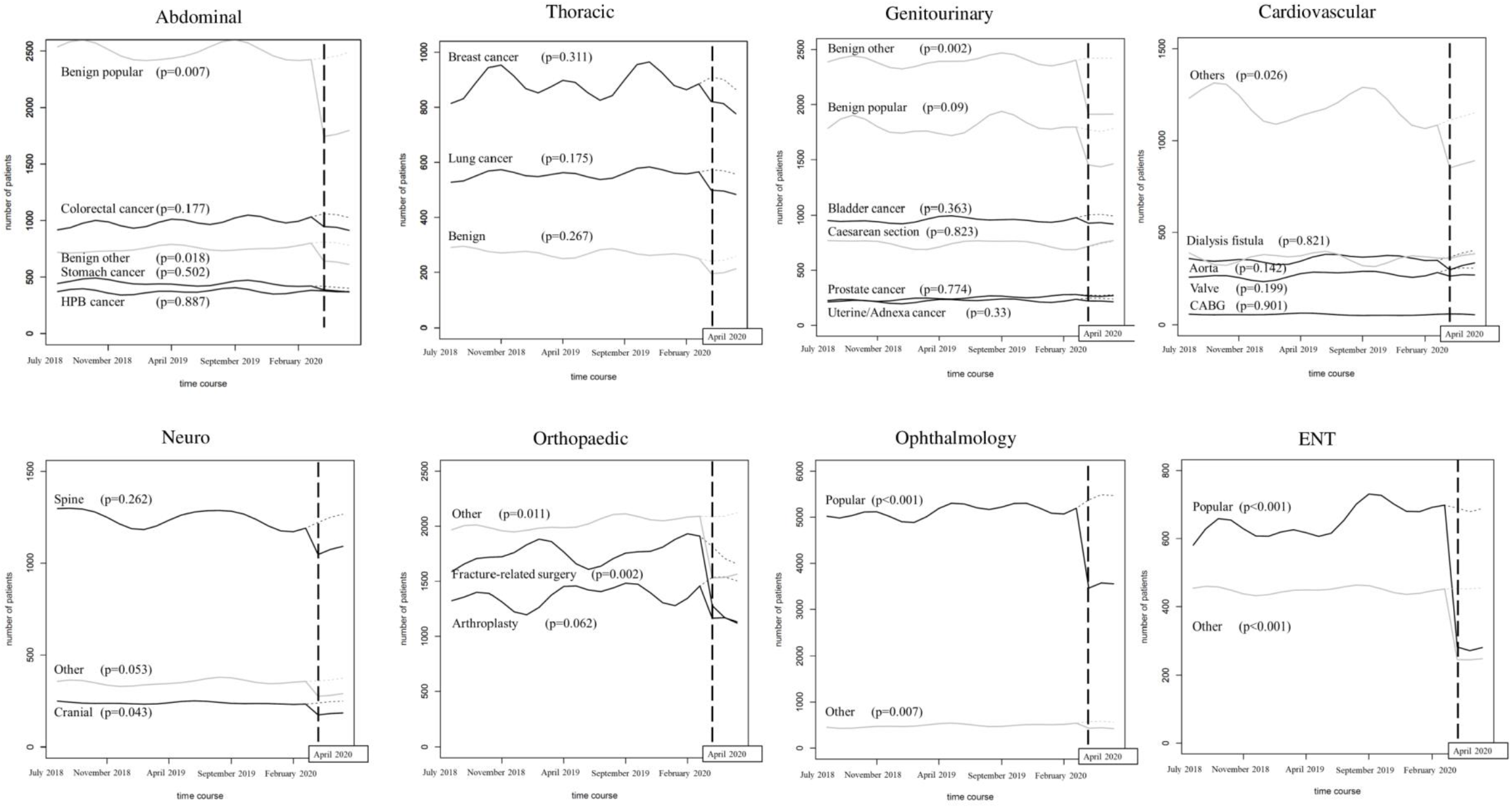
Monthly discharge volume of each surgical category over time with interrupted time series analysis including segmented regressions. ENT, Ear/Nose/Throat

### 3.3. Thoracic surgery

The trend was gradually decreasing in all type of surgeries. Only prior year comparison of benign surgeries was lower from March (supplemental Figure 2, 84.9%). ITS analysis showed that the reduction of every type of surgical volume was not statistically significant (Figure 4: lung cancer, p = 0.175; breast cancer, p = 0.311, benign surgery, p = 0.267).

### 3.4. Genitourinary surgery

The number of oncological surgeries and caesarean sections during March, April, and May in 2020 did not show a significant decrease compared to the same months in the previous year. In terms of benign surgeries, there was a slight downward trend in April and May (supplemental Figure 3: benign popular, 92.4% in April and 75.9% in May; benign other, 91.7% in April and 70.9% in May). ITS analysis showed a statistically significant effect of COVID-19 only on other benign surgeries (Figure 4, p = 0.005).

### 3.5. Cardiovascular surgery

The number of surgeries for valve, aorta, CABG, and others gradually decreased from April to a minimum in May (supplemental Figure 4: valve, 74.9%; aorta, 83.4%; CABG, 71.2%; dialysis fistula, 81.9%, other, 62.6%). ITS analysis showed that only other types of surgery were statistically significant (Figure 4, p = 0.026).

### 3.6. Neurosurgery

All type of surgeries had low numbers compared to the previous year in April and May. Especially, May had the lowest percentage (supplemental Figure 5: cranial, 68.2%; spine, 71.8%; other, 60.3%). On the other hand, only ITS analysis of intracranial surgery showed a statistically significant decline (Figure 4; cranial, p = 0.043; spine, p = 0.262; other, p = 0.053).

### 3.7. Orthopaedic surgery

All categories gradually decreased from April. Fracture-related surgery in May was the lowest among all comparisons in this pandemic period (supplemental Figure 6: fracture-related surgery, 60.6%; spine, 65.2%; other, 62.1%). All surgeries except arthroplasty were statistically significant according to the ITS analysis (Figure 4: fracture-related surgery, p = 0.002; arthroplasty, p = 0.062; other, p = 0.011).

### 3.8. Ophthalmologic surgery

Most forms of ophthalmic surgery were in the category of popular surgery. The year-over-year trend of both popular and other type of surgery decreased from April, especially in May (supplemental Figure 7: popular, 51.0%; other, 69.1%). Decline in both types of surgery was significantly affected by COVID-19 (Figure 4: popular, p < 0.001; other, p = 0.007).

### 3.9. Ear/Nose/Throat surgery

The trends of both popular and other types of surgery per month were similar, with dramatic decreases in May (supplemental Figure 8: popular, 21.6%; other, 35.0%). Showing the same trend as in the descriptive analysis, ITS analysis showed statistically significant decrease in both surgical types (Figure 4: popular, p < 0.001; other, p < 0.001).

## 4. Discussion

Our study showed the trends of scheduled admission with surgery and important changes in surgical volume during the COVID-19 pandemic in Japan. The main findings of our study were as follows: first, we found the most affected practice was ENT surgery according to the year-over-year trend; second, even within the same category, benign surgeries that were considered non-critical tended to be significantly affected by the JSS’s announcement of surgical triage due to COVID-19.

First, the decline of oncological surgeries for gastrointestinal, HPB, lung, breast, and genitourinary cancer was not significant in the ITS analysis, even though there were reductions when compared to the same period of the last year. For patients who had nonfatal or did not require urgent medical intervention, JSS recommended postponement or performing surgery cautiously under appropriate infection control.^5^ Because of the risk of cancer-progression, prioritisation of procedures should consider available evidence on time for surgery and oncologic outcomes.^9,19,20^ The American College of Surgeons also recommended that surgery for low-risk cancer should be deferred but should not be postponed for most other cancers.^21^ For example, our result for gastrointestinal cancer could reflect the recommendation because we did not include endoscopic procedures, which were usually performed for early stage cancer outside the operating room. However, our result for prostate cancer showed no downward trend, which was opposite to urological triage recommendations.^22^ This was probably because of the relatively affordability of hospital beds recruited in this study and the patients’ wishes to perform the surgery as scheduled. In terms of the benign abdominal, thoracic, and genitourinary surgeries, all but thoracic and popular-genitourinary decreased with p < 0.05 in ITS analysis. In our dataset, the most popular thoracic benign surgery was the video-assisted thoracoscopic surgery for the pneumothorax, and the most popular benign genitourinary was stone surgery. Both surgeries have a high priority and are basically recommended to be performed without delay.^5,9,22^

Second, within cardiovascular surgery types, only other types of surgery obviously decreased. During the COVID-19 pandemic, management of ICU beds and ventilators is important. There is also concern about a remarkable reduction in blood donation because many cardiovascular surgeries require perioperative transfusion.^23,24^ Considering the risk of COVID-19 infection and conservation of medical resources, certain surgical restrictions, such as delaying of surgery for stable patients or transferring a patient to near-by hospital, are recommended for cardiac and aortic surgery by the Japanese Society for Cardiovascular Surgery, as in western countries.^25-27^ However, cardiovascular surgery is thought to be essential because of the risk of worsening or even death caused by surgical delay. For example, progression from asymptomatic to symptomatic (typically chest pain or shortness of breath) with valvular disease or aortic aneurysm can increase the risk of surgery, or even worse, make surgery impossible. Similarly, if the appropriate timing of surgery for dialysis fistula is missed, patient’s condition will become irreversible because of kidney failure. When medical resources, such as hospital beds, staff, and ventilators can be afforded to a certain extent, many cardiovascular surgeries should not be unnecessarily postponed, and such a trend was also observed in this study.

Third, within neurosurgery, although every type declined compared to the same period in the previous year, only intracranial surgery was significant with p < 0.05. According to a report by Johns Hopkins University, the decrease rate of cranial surgery cases was largest in the department of neurosurgery.^28^ Triage of elective neurosurgery was basically judged by the symptoms, such as any signs of life-, limb- or vision-threatening conditions.^29^ The Japan Neurosurgical Society also announced similar statements with only an additional alert about endoscopic surgery.^30^ In this research, we chose surgeries for brain tumour, intracranial haemorrhage, and intracranial vascular as cranial surgery based on their popularity in the NDB. Because patients who underwent these surgeries after scheduled admission might possibly have had trivial symptoms, our results could have been affected by such selection effects. Spine surgery also decreased, although the ITS analysis was not significant. The Japan Neurosurgical Society announced that spine surgery is intermediate or high priory according to the symptoms.^30^ Patients who received spinal injury often suffered from paralysis, which leads to decline in activities of daily living (ADL) or quality of life, and the complaints of symptoms had possibly become more pronounced, which might have led to our results.

Lastly, surgeries associated with quality of life such as orthopaedic, ophthalmologic, and ENT surgeries were focused on. Traumatic surgery is one that requires minimising preoperative delay and LOS despite a patient’s COVID-19 status at each hospital,^9^ although whether surgery is performed with optimal timing or not should be decided in each hospital based on restrictions on elective resources and procedures.^31^ Considering the necessity of traumatic surgery such as fracture, surgical volume might have declined because of the state of emergency declared by the government that led to people voluntarily refraining from going out. Some arthroplasty cases were performed for fracture patients, although most of them were for osteoarthritis. This might be because we selected only scheduled admission patients. Osteoarthritis decreases ADL regardless of there being a state of emergency, which could be related to our results. The Japanese Ophthalmological Society and Oto-Rhino-Laryngological Society of Japan recommended cancellation or postponement of most elective surgeries because of the high probability of the COVID-19 virus being present in the nasal cavity, pharynx, and lacrimal fluid.^32,33^ Moreover, both ophthalmologists and otolaryngologists are a high-risk category of COVID-19 infection because their daily outpatient and emergent patients have a high volume. In our study, the reason for the dramatic decrease in the surgical volume in otolaryngology and ophthalmology compared to other surgical categories might be that patients themselves were reluctant to be seen and that physicians in these two departments were strictly triaged by urgency.

Our study has several limitations. First, our target hospitals were 254 QIP participant that voluntarily provided DPC data. This study population may not have been comprehensive enough to be representative of the current situation in Japan because there are more than 8000 hospitals in Japan. In this study, participant hospitals had a variety of backgrounds and corona-positive patients were admitted in at least about half of the hospitals. We believe that selection bias was not very significant, and this result was important for understanding the future course of the pandemic. Second, we did not consider whether there was a trend for surgery originally performed in an inpatient setting for ambulatory surgery because of the COVID-19 pandemic. The Centers for Disease Control and Prevention has recommended that some inpatient surgeries should be shifted to ambulatory surgery when possible and it is safe to do so, and the Ambulatory Surgery Center Association also cited the recommendation because of the risk of infection for medical staff and patients and the efficiency of medical resources.^34^ Ambulatory surgery centres need to be prepared for the possibility of performing surgeries such as trauma and backlog surgeries that have never been experienced before depending on how the COVID-19 pandemic develops in the future. Finally, the ITS analysis was performed with DPC data based on the discharge date. The state of emergency declared by the government and surgical triage by each hospital for the COVID-19 pandemic would affect the date of admissions, not discharges. However, we could only acquire the data of patients who were discharged by June 30, 2020. That is why we separated the target population for descriptive analysis from that of ITS analysis. To support our results, we added information about the median and inter-quartile range of LOS that were varied from surgeries to the supplemental tables for ITS analysis.

In conclusion, we demonstrated and validated the trend of surgical volume reduction in Japan, by applying ITS analyses. The most affected practice was ENT surgery, especially popular surgeries, with declines of about 80%. Benign surgeries, except critical procedures, tended to be significantly affected by COVID-19. Future research should examine how hospitals recovered efficiently from the backlog of surgeries during this first wave of the pandemic, using more recent administrative data, to prepare for the next possible pandemic wave after winter 2020.

## Data Availability

Due to the sensitive nature of the data derived from a government database, raw data would not be shared.
Data not available / The data that has been used is confidential

## Acknowledgments

We thank all the staff members and all the participating acute care hospitals. Our database using in this research cannot be available publicly because of their nature (the data of Japanese government).

## Figure legends

Supplementary Fig. 1. Year-over-year trend in number by major surgical type within abdominal surgeries

Supplementary Fig. 2. Year-over-year trend in number by major surgical type within thoracic surgeries

Supplementary Fig. 3. Year-over-year trend in number by major surgical type within genitourinary surgeries

Supplementary Fig. 4. Year-over-year trend in number by major surgical type within cardiovascular surgeries

Supplementary Fig. 5. Year-over-year trend in number by major surgical type within neurosurgeries

Supplementary Fig. 6. Year-over-year trend in number by major surgical type within orthopaedic surgeries

Supplementary Fig. 7. Year-over-year trend in number by major surgical type within ophthalmologic surgeries

Supplementary Fig. 8. Year-over-year trend in number by major surgical type within Ear/Nose/Throat surgeries

