## supplemental tables for "IMPACT OF THE CORONAVIRUS DISEASE 2019 PANDEMIC ON SURGICAL VOLUME IN JAPAN: A COHORT STUDY USING ADMINISTRATIVE DATA"

Supplemental Table 1

| **Abdominal** | Oncology 　Stomach, K6552, K655-22, K655-42, K655-52, K657-22, K6572 　Colorectal, K740, K7193, K719-3; HPB, K695, K70 [2-4], K67 [5-7] Benign 　Popular, K633, K634, K672, K718 　Others, K63 [6-9], K64 [0-6,8,9], K652, K659, K66 [0-3,6,7,9], K67 [0,3,4,9], K680, K69 [0,2-4,6,8], K70 [0,1,5,6], K71 [2-4,6,7], K72 [0,6,9], K73 [0-4,6], K74 [1,2], K75 [0-3] |
| --- | --- |
| **Thoracic** | Oncology: Lung, K514; Breast: K476 Benign: K47 [4,5,8], K48 [0-3,5-8], K50 [1-3,7-9], K51 [1,3,5-9] |
| **Genitourinary** | Oncology 　Bladder, K803; Prostate, K843; Uterine/Adnexa, K879, K889 Benign 　Popular, K78 [12], K798, K87 [67], K888  　Others, K75 [45,7-9], K76, K77 [02,5-9], K78 [4-9], K79 [0-7,9], K80 [124,6-9], K81 [0125689], K82, K83 [02,4-9], K8 [5,6], K87 [1-3,8], K88 [1-7], K890, K90 [3-6], K912  Caesarean section, K898 |
| **Cardiovascular** | Valve, K55 [4-6]; Aortic, K560, K561; CABG: K552;  Dialysis fistula: K612 [1-3], K610-3  Others: K55 [3678], K56 [2-9], K5 [78], K59 [0-4], K60 [3-5,7-9], K610 [1-5], K610- [245], K61 [4579], K623 |
| **Neurosurgical** | Cranial, K16 [4,7,9], K17 [0,1,6,7]; Spine, K142, K134  Others: K128, K13 [13], K13 [5-9], K14 [01], K14 [3-9], K15, K16 [0-3], K16 [5-8], K17 [2-5], K179, K18 [0-8], K19 [0-8] |
| **Orthopaedic** | Fracture, K04 [568]; Arthroplasty, K08 [12]  Others: K02 [5689], K03 [01345789], K04 [039], K05 [0-9], K06 [0,3-9], K07 [0234,6-9], K08 [0,3-8], K09 [34,6-9], K10, K11 [23689], K12 [01456] |
| **Ophthalmology** | Popular, K21 [7-9], K280, K282  Others, K199, K20, K21 [1-6], K2 [2-7], K28 [14] |
| **Ear/Nose/Throat** | Popular, K319, K340- [3-7], K377  Others, K28 [5-9], K29, K3 [0-3], K3401, K34 [1-7], K3 [56], K37 [0-6], K37 [89], K38 [0-5], K38 [7-9], K39, K40 [0-3] |

HPB, hepato‑pancreatic‑biliary

Supplemental Table 2

Raw data of year-over-year trend in the number of surgeries by the eight categories

|  |  | Jul | Aug | Sep | Oct | Nov | Dec | Jan | Feb | Mar | Apr | May | Jun |
| --- | --- | --- | --- | --- | --- | --- | --- | --- | --- | --- | --- | --- | --- |
| Abdominal | July 2018 to June 2019 | 5147 | 5230 | 4568 | 5398 | 5146 | 4519 | 5153 | 4970 | 5062 | 4806 | 4610 | 3683 |
|  | July 2019 to June 2020 | 5380 | 5106 | 5003 | 5330 | 4979 | 4782 | 5076 | 4869 | 5178 | 4317 | 3157 | 3081 |
| Thoracic | July 2018 to June 2019 | 1755 | 1710 | 1615 | 1878 | 1748 | 1627 | 1749 | 1647 | 1640 | 1722 | 1523 | 1321 |
|  | July 2019 to June 2020 | 1789 | 1612 | 1660 | 1793 | 1750 | 1731 | 1790 | 1658 | 1795 | 1599 | 1307 | 1160 |
| Genitourinary | July 2018 to June 2019 | 6348 | 6635 | 5553 | 6650 | 5936 | 5678 | 6093 | 5953 | 5975 | 5831 | 5764 | 5221 |
|  | July 2019 to June 2020 | 6669 | 6342 | 5990 | 6431 | 5860 | 6103 | 6195 | 5791 | 6365 | 5644 | 4530 | 4424 |
| Cardiovascular | July 2018 to June 2019 | 2434 | 2260 | 2145 | 2310 | 2333 | 1904 | 2194 | 2099 | 2087 | 2139 | 2006 | 1552 |
|  | July 2019 to June 2020 | 2535 | 2180 | 2194 | 2277 | 2281 | 2104 | 2277 | 2060 | 2167 | 1943 | 1428 | 1389 |
| Neurosurgical | July 2018 to June 2019 | 1988 | 1883 | 1645 | 1972 | 1845 | 1612 | 1794 | 1810 | 1736 | 1711 | 1624 | 1022 |
|  | July 2019 to June 2020 | 2072 | 1909 | 1770 | 1804 | 1779 | 1710 | 1815 | 1721 | 1854 | 1578 | 1122 | 937 |
| Orthopaedic | July 2018 to June 2019 | 5068 | 5269 | 4417 | 5284 | 5173 | 4507 | 5289 | 5194 | 5392 | 4928 | 4454 | 3040 |
|  | July 2019 to June 2020 | 5649 | 5189 | 4865 | 5433 | 5234 | 4960 | 5498 | 5194 | 5600 | 4035 | 2781 | 2544 |
| Ophthalmologic | July 2018 to June 2019 | 5921 | 5314 | 4906 | 6038 | 6016 | 4747 | 5416 | 5710 | 5554 | 5314 | 5677 | 5721 |
|  | July 2019 to June 2020 | 6514 | 5171 | 5578 | 5768 | 6026 | 5247 | 5860 | 5376 | 6094 | 4216 | 2952 | 4175 |
| Ear/Nose/Throat | July 2018 to June 2019 | 1066 | 1174 | 961 | 1116 | 1129 | 954 | 1059 | 1111 | 1047 | 1082 | 959 | 892 |
|  | July 2019 to June 2020 | 1188 | 1165 | 1202 | 1210 | 1071 | 1120 | 1129 | 1090 | 1136 | 580 | 262 | 528 |

Supplemental Table 3

Baseline demographics of the cases within abdominal surgery group for interrupted time series analysis

|  | Stomach cancer | |  | Colorectal cancer | |  | HPB cancer | |  |
| --- | --- | --- | --- | --- | --- | --- | --- | --- | --- |
|  | Before | After | P-value | Before | After | P-value | Before | After | P-value |
| n | 9,369 | 1,133 |  | 20696 | 2800 |  | 7819 | 1116 |  |
| Sex = male, n (%) | 6,535 (69.8) | 841 (74.2) | 0.002 | 11810 (57.1) | 1628 (58.1) | 0.288 | 5056 (64.7) | 730 (65.4) | 0.648 |
| Age (mean (SD)) | 71.68 (10.19) | 72.11 (10.16) | 0.178 | 70.74 (11.12) | 70.69 (11.35) | 0.843 | 70.85 (9.56) | 71.41 (9.62) | 0.07 |
| CCI > 2, n (%) | 1,336 (14.3) | 156 (13.8) | 0.688 | 3637 (17.6) | 464 (16.6) | 0.199 | 1736 (22.2) | 240 (21.5) | 0.627 |
| EI > 5, n (%) | 1,740 (18.6) | 177 (15.6) | 0.017 | 4154 (20.1) | 570 (20.4) | 0.742 | 2481 (31.7) | 307 (27.5) | 0.005 |
| LOS (median [IQR]) | 16 [13, 22] | 16 [12, 21] | 0.179 | 15 [12, 21] | 14 [11, 20] | <0.001 | 17 [12, 28] | 16 [12, 26] | 0.001 |
|  | Benign (popular) | |  | Benign (other) | |  | Benign surgery | |  |
|  | Before | After | P-value | Before | After | P-value | Before | After | P-value |
| n | 52434 | 5305 |  | 15747 | 1887 |  | 68181 | 7192 |  |
| Sex = male, n (%) | 36082 (68.8) | 3822 (72.0) | <0.001 | 8574 (54.4) | 1013 (53.7) | 0.544 | 44656 (65.5) | 4835 (67.2) | 0.003 |
| Age (mean (SD)) | 64.87 (15.22) | 65.22 (15.14) | 0.114 | 64.27 (16.07) | 64.55 (16.01) | 0.473 | 64.73 (15.42) | 65.04 (15.37) | 0.105 |
| CCI > 2, n (%) | 2323 (4.4) | 272 (5.1) | 0.021 | 2108 (13.4) | 263 (13.9) | 0.531 | 4431 (6.5) | 535 (7.4) | 0.002 |
| EI > 5, n (%) | 4719 (9.0) | 528 (10.0) | 0.023 | 2560 (16.3) | 320 (17.0) | 0.456 | 7279 (10.7) | 848 (11.8) | 0.004 |
| LOS (median [IQR]) | 5 [4, 6] | 5 [4, 6] | 0.599 | 11 [7, 16] | 11 [8, 17] | 0.002 | 17 [12, 28] | 16 [12, 26] | 0.001 |

HPB, hepato‑pancreatic‑biliary; LOS, length of stay; IQR, inter-quartile range

Supplemental Table 4

Baseline demographics of the cases within thoracic surgery group for interrupted time series analysis

|  | Lung cancer | |  | Breast cancer | |  | Benign | |  |
| --- | --- | --- | --- | --- | --- | --- | --- | --- | --- |
|  | Before | After | P-value | Before | After | P-value | Before | After | P-value |
| n | 11699 | 1478 |  | 18627 | 2411 |  | 5689 | 607 |  |
| Sex = male, n (%) | 7102 (60.7) | 883 (59.7) | 0.493 | 95 (0.5) | 14 (0.6) | 0.761 | 2675 (47.0) | 312 (51.4) | 0.044 |
| Age (mean (SD)) | 70.46 (9.31) | 70.87 (9.55) | 0.11 | 62.30 (14.08) | 61.84 (14.18) | 0.137 | 54.32 (20.03) | 56.05 (19.36) | 0.042 |
| CCI > 2, n (%) | 2020 (17.3) | 321 (21.7) | <0.001 | 1604 (8.6) | 210 (8.7) | 0.901 | 424 (7.5) | 49 (8.1) | 0.639 |
| EI > 5, n (%) | 2176 (18.6) | 336 (22.7) | <0.001 | 2038 (10.9) | 257 (10.7) | 0.702 | 542 (9.5) | 62 (10.2) | 0.636 |
| LOS (median [IQR]) | 10 [8, 13] | 10 [8, 13] | 0.495 | 8 [6, 10] | 8 [6, 10] | 0.021 | 6 [4, 9] | 7 [4, 10] | 0.042 |

LOS, length of stay; IQR, inter-quartile range

Supplemental Table 5

Baseline demographics of the cases within genitourinary surgery group for interrupted time series analysis

|  | Bladder cancer | |  | Prostate cancer | |  | Uterine/Adnexa cancer | | | |  |
| --- | --- | --- | --- | --- | --- | --- | --- | --- | --- | --- | --- |
|  | Before | After | P-value | Before | After | P-value | Before | | After | | P-value |
| n | 20,046 | 2,781 |  | 5187 | 799 |  | 4651 | | 655 | |  |
| Sex = male, n (%) | 15993 (79.8) | 2245 (80.7) | 0.254 | 5187 (100.0) | 799 (100.0) | NA | 4651 (100.0) | | 655 (100.0) | | NA |
| Age (mean (SD)) | 74.15 (10.06) | 74.41 (9.76) | 0.202 | 69.44 (5.85) | 69.22 (6.47) | 0.331 | 58.96 (13.20) | | 58.46 (13.00) | | 0.364 |
| CCI > 2, n (%) | 1946 (9.7) | 311 (11.2) | 0.016 | 195 (3.8) | 22 (2.8) | 0.189 | 424 (9.1) | | 61 (9.3) | | 0.927 |
| EI > 5, n (%) | 2314 (11.5) | 350 (12.6) | 0.116 | 331 (6.4) | 41 (5.1) | 0.199 | 473 (10.2) | | 72 (11.0) | | 0.562 |
| LOS (median [IQR]) | 6 [5, 8] | 6 [5, 8] | 0.424 | 11 [10, 12] | 10 [9, 12] | 0.011 | 11 [9, 15] | | 10 [9, 13] | | <0.001 |
|  | Benign (popular) | |  | Benign (other) | |  | Caesarean delivery | | | |  |
|  | Before | After | P-value | Before | After | P-value | Before | | After | | P-value |
| n | 37981 | 4347 | 0.078 | 50307 | 5737 |  | 15526 | | 2236 | |  |
| Sex = male, n (%) | 8795 (23.2) | 1059 (24.4) | 0.019 | 19533 (38.8) | 2304 (40.2) | 0.052 | NA | | NA | | NA |
| Age (mean (SD)) | 52.94 (15.83) | 53.53 (15.93) | 1 | 56.27 (17.60) | 56.53 (17.70) | 0.281 | 33.51 (5.16) | | 33.49 (5.16) | | 0.887 |
| CCI > 2, n (%) | 645 (1.7) | 74 (1.7) | 0.028 | 1440 (2.9) | 190 (3.3) | 0.06 | 5 (0.0) | | 1 (0.0) | | 1 |
| EC > 5, n (%) | 1425 (3.8) | 193 (4.4) | <0.001 | 2178 (4.3) | 281 (4.9) | 0.05 | 96 (0.6) | | 19 (0.8) | | 0.257 |
| LOS (median [IQR]) | 6 [5, 8] | 6 [5, 8] | 0.452 | 5 [3, 8] | 6 [3, 9] | 0.008 | 9 [8, 10] | | 9 [8, 10] | | 0.001 |

CCI, Charlson comorbidity index; EC, Elixhauser comorbidity LOS, length of stay; IQR, inter-quartile range

Supplemental Table 6

Baseline demographics of the cases within cardiovascular surgery group for interrupted time series analysis

|  | Valve | | | |  | | Aorta | | | |  | | CABG | | | | | | | | |  |
| --- | --- | --- | --- | --- | --- | --- | --- | --- | --- | --- | --- | --- | --- | --- | --- | --- | --- | --- | --- | --- | --- | --- |
|  | Before | | After | | P-value | | Before | | After | | P-value | | Before | | | | After | | | | | P-value |
| n | 5592 | | 800 | |  | | 7463 | | 950 | |  | | 1170 | | | | 173 | | | | |  |
| Sex = male, n (%) | 2685 (48.0) | | 409 (51.1) | | 0.108 | | 5926 (79.4) | | 732 (77.1) | | 0.101 | | 919 (78.5) | | | | 138 (79.8) | | | | | 0.79 |
| Age (mean (SD)) | 74.67 (12.00) | | 74.99 (11.69) | | 0.472 | | 73.83 (9.80) | | 73.56 (9.92) | | 0.428 | | 69.69 (9.67) | | | | 69.22 (10.37) | | | | | 0.556 |
| CCI > 2, n (%) | 1117 (20.0) | | 156 (19.5) | | 0.789 | | 1217 (16.3) | | 152 (16.0) | | 0.845 | | 409 (35.0) | | | | 62 (35.8) | | | | | 0.888 |
| EC > 5, n (%) | 3166 (56.6) | | 472 (59.0) | | 0.217 | | 2319 (31.1) | | 290 (30.5) | | 0.76 | | 560 (47.9) | | | | 92 (53.2) | | | | | 0.221 |
| LOS (median [IQR]) | 19 [14, 28] | | 18 [12, 26] | | <0.001 | | 14 [10, 21] | | 13 [10, 19] | | 0.03 | | 24.50[19, 33] | | | | 25 [19, 34] | | | | | 0.82 |
|  | Shunt | | | |  | | Others | | | |  | | |  | | | |  | | |  | |
|  | Before | | After | | P-value | | Before | | After | | P-value | |  | | |  | | | |  | | |
| n | 7537 | | 1116 | |  | | 24857 | | 2602 | |  | |  | | |  | | | |  | | |
| Sex = male, n (%) | 5187 (68.8) | | 770 (69.0) | | 0.933 | | 14703 (59.2) | | 1709 (65.7) | | <0.001 | |  | | |  | | | |  | | |
| Age (mean (SD)) | 69.13 (12.82) | | 69.39 (12.88) | | 0.532 | | 70.40 (11.89) | | 71.03 (11.56) | | 0.01 | |  | | |  | | | |  | | |
| CCI > 2, n (%) | 1737 (23.0) | | 240 (21.5) | | 0.269 | | 3865 (15.5) | | 460 (17.7) | | 0.005 | |  | | |  | | | |  | | |
| EC > 5, n (%) | 1802 (23.9) | | 276 (24.7) | | 0.574 | | 7850 (31.6) | | 902 (34.7) | | 0.001 | |  | | |  | | | |  | | |
| LOS (median [IQR]) | 5 [3, 15] | | 5 [3, 14] | | 0.705 | | 7 [3, 11] | | 8 [4, 13] | | <0.001 | |  | | |  | | | |  | | |

CCI, Charlson comorbidity index; EC, Elixhauser comorbidity LOS, length of stay; IQR, inter-quartile range

Supplemental Table 7

Baseline demographics of the cases within neuro surgery group for interrupted time series analysis

|  | Cranial | |  | Spine | |  | Others | |  |
| --- | --- | --- | --- | --- | --- | --- | --- | --- | --- |
|  | Before | After | P-value | Before | After | P-value | Before | After | P-value |
| n | 4997 | 538 |  | 26083 | 3213 |  | 7368 | 843 |  |
| Sex = male, n (%) | 2193 (43.9) | 233 (43.3) | 0.833 | 15430 (59.2) | 1993 (62.0) | 0.002 | 4012 (54.5) | 481 (57.1) | 0.16 |
| Age (mean (SD)) | 65.24 (13.64) | 65.83 (14.20) | 0.344 | 67.12 (14.39) | 65.99 (14.83) | <0.001 | 59.96 (18.43) | 60.46 (18.05) | 0.458 |
| CCI > 2, n (%) | 410 (8.2) | 41 (7.6) | 0.698 | 1579 (6.1) | 216 (6.7) | 0.146 | 296 (4.0) | 53 (6.3) | 0.003 |
| EC > 5, n (%) | 1158 (23.2) | 133 (24.7) | 0.452 | 2716 (10.4) | 322 (10.0) | 0.512 | 979 (13.3) | 125 (14.8) | 0.234 |
| LOS (median [IQR]) | 15 [11, 22] | 15[11, 22.75] | 0.879 | 16 [12, 23] | 15 [11, 21] | <0.001 | 11 [4, 17] | 11 [4, 18] | 0.815 |

CCI, Charlson comorbidity index; EC, Elixhauser comorbidity LOS, length of stay; IQR, inter-quartile range

Supplemental Table 8

Baseline demographics of the cases within orthopaedic surgery group for interrupted time series analysis

|  | Fracture | |  | Arthroplasty | |  | Others | |  |
| --- | --- | --- | --- | --- | --- | --- | --- | --- | --- |
|  | Before | After | P-value | Before | After | P-value | Before | After | P-value |
| n | 36931 | 3569 |  | 28749 | 3465 |  | 42535 | 4628 |  |
| Sex = male, n (%) | 17266 (46.8) | 1664 (46.6) | 0.897 | 5963 (20.7) | 773 (22.3) | 0.034 | 21756 (51.1) | 2437 (52.7) | 0.053 |
| Age (mean (SD)) | 56.07 (19.99) | 58.15 (19.00) | <0.001 | 72.59 (9.80) | 72.84 (9.62) | 0.149 | 54.64 (19.81) | 56.53 (18.54) | <0.001 |
| CCI > 2, n (%) | 857 (2.3) | 110 (3.1) | 0.005 | 1072 (3.7) | 152 (4.4) | 0.062 | 1512 (3.6) | 214 (4.6) | <0.001 |
| EC > 5, n (%) | 1840 (5.0) | 209 (5.9) | 0.025 | 2248 (7.8) | 294 (8.5) | 0.18 | 2168 (5.1) | 283 (6.1) | 0.003 |
| LOS (median [IQR]) | 4 [3, 8] | 4 [3, 9] | <0.001 | 23 [18, 32] | 23 [17, 32] | 0.025 | 9 [4, 20] | 10 [4, 23] | <0.001 |

CCI, Charlson comorbidity index; EC, Elixhauser comorbidity LOS, length of stay; IQR, inter-quartile range

Supplemental Table 9

Baseline demographics of the cases within ophthalmologic surgery group for interrupted time series analysis

|  | Popular | |  | Other | |  |
| --- | --- | --- | --- | --- | --- | --- |
|  | Before | After | P-value | Before | After | P-value |
| n | 107647 | 10591 |  | 10270 | 1299 |  |
| Sex = male, n (%) | 46522 (43.2) | 5060 (47.8) | <0.001 | 5497 (53.5) | 704 (54.2) | 0.669 |
| Age (mean (SD)) | 74.50 (9.90) | 73.82 (10.08) | <0.001 | 66.39 (16.59) | 66.95 (16.09) | 0.246 |
| CCI > 2, n (%) | 3842 (3.6) | 448 (4.2) | 0.001 | 294 (2.9) | 44 (3.4) | 0.332 |
| EC > 5, n (%) | 4204 (3.9) | 515 (4.9) | <0.001 | 306 (3.0) | 43 (3.3) | 0.568 |
| LOS (median [IQR]) | 3 [2, 4] | 3 [2, 4] | 0.001 | 5 [3, 9] | 5 [3, 8] | 0.035 |

CCI, Charlson comorbidity index; EC, Elixhauser comorbidity LOS, length of stay; IQR, inter-quartile range

Supplemental Table 10

Baseline demographics of the cases within Ear/Nose/Throat surgery group for interrupted time series analysis

|  | Popular | |  | Others | |  |
| --- | --- | --- | --- | --- | --- | --- |
|  | Before | After | P-value | Before | After | P-value |
| n | 13714 | 836 |  | 9412 | 739 |  |
| Sex = male, n (%) | 7612 (55.5) | 487 (58.3) | 0.129 | 6278 (66.7) | 513 (69.4) | 0.141 |
| Age (mean (SD)) | 47.38 (19.40) | 48.98 (18.83) | 0.02 | 53.08 (20.22) | 57.45 (20.01) | <0.001 |
| CCI > 2, n (%) | 142 (1.0) | 2 (0.2) | 0.038 | 549 (5.8) | 63 (8.5) | 0.004 |
| EC > 5, n (%) | 372 (2.7) | 27 (3.2) | 0.436 | 793 (8.4) | 91 (12.3) | <0.001 |
| LOS (median [IQR]) | 8 [6, 8] | 7 [6, 9] | 0.194 | 6 [3, 8] | 6 [3, 9.50] | <0.001 |

CCI, Charlson comorbidity index; EC, Elixhauser comorbidity LOS, length of stay; IQR, inter-quartile range
