## Supplementary figures and images for "IMPACT OF THE CORONAVIRUS DISEASE 2019 PANDEMIC ON SURGICAL VOLUME IN JAPAN: A COHORT STUDY USING ADMINISTRATIVE DATA"

### supplemental figure1

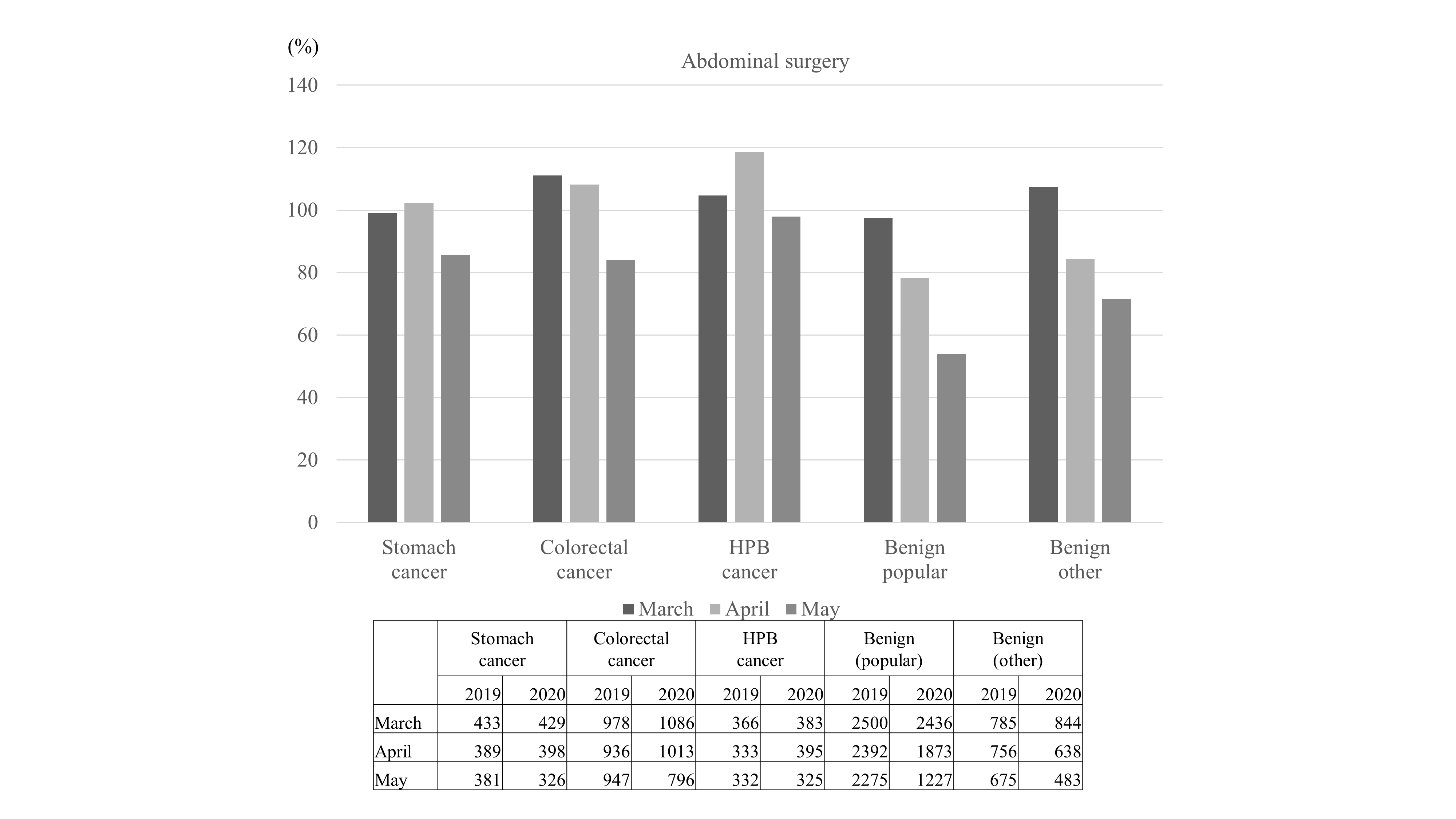

### supplemental figure2

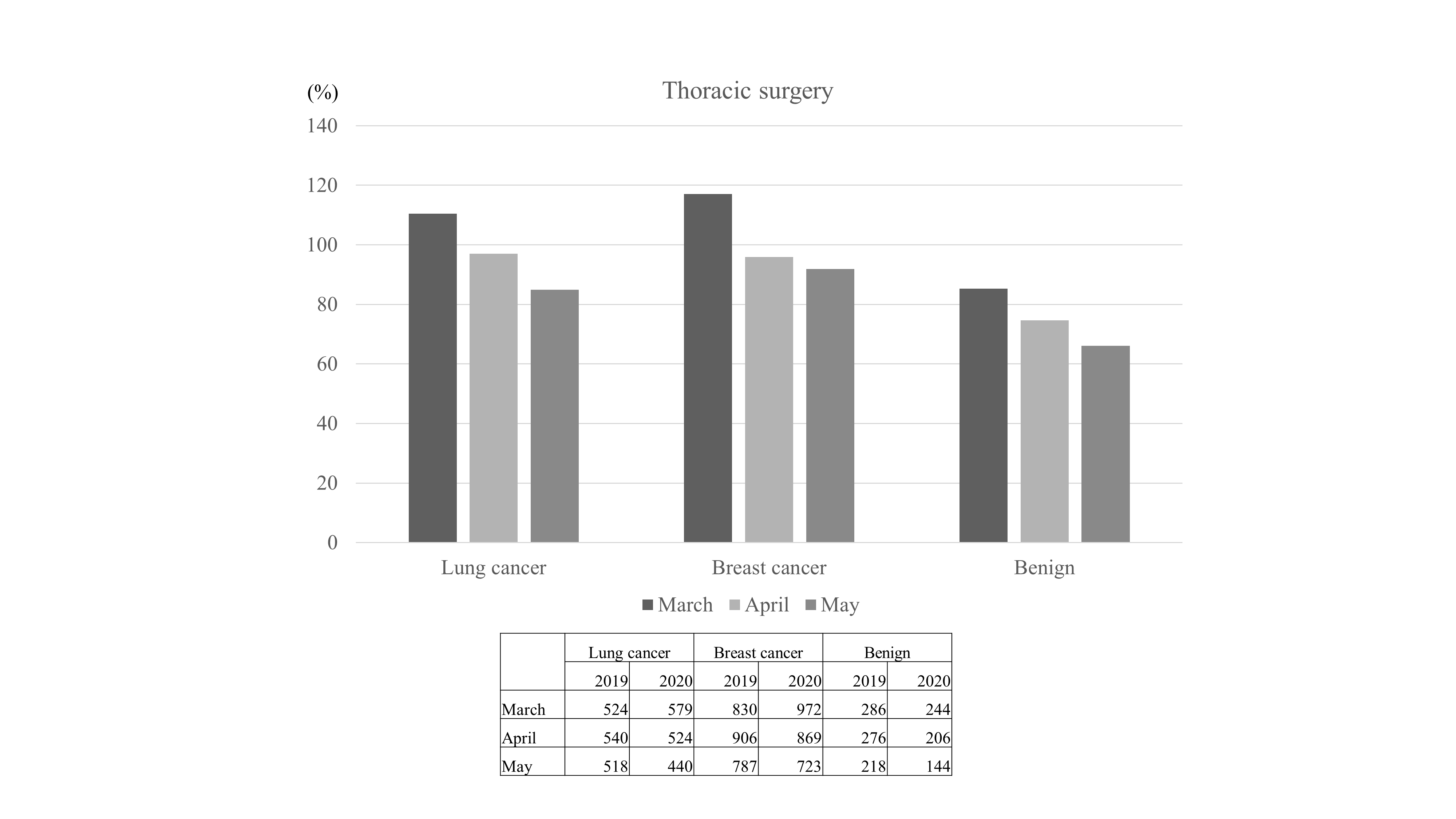

### supplemental figure3

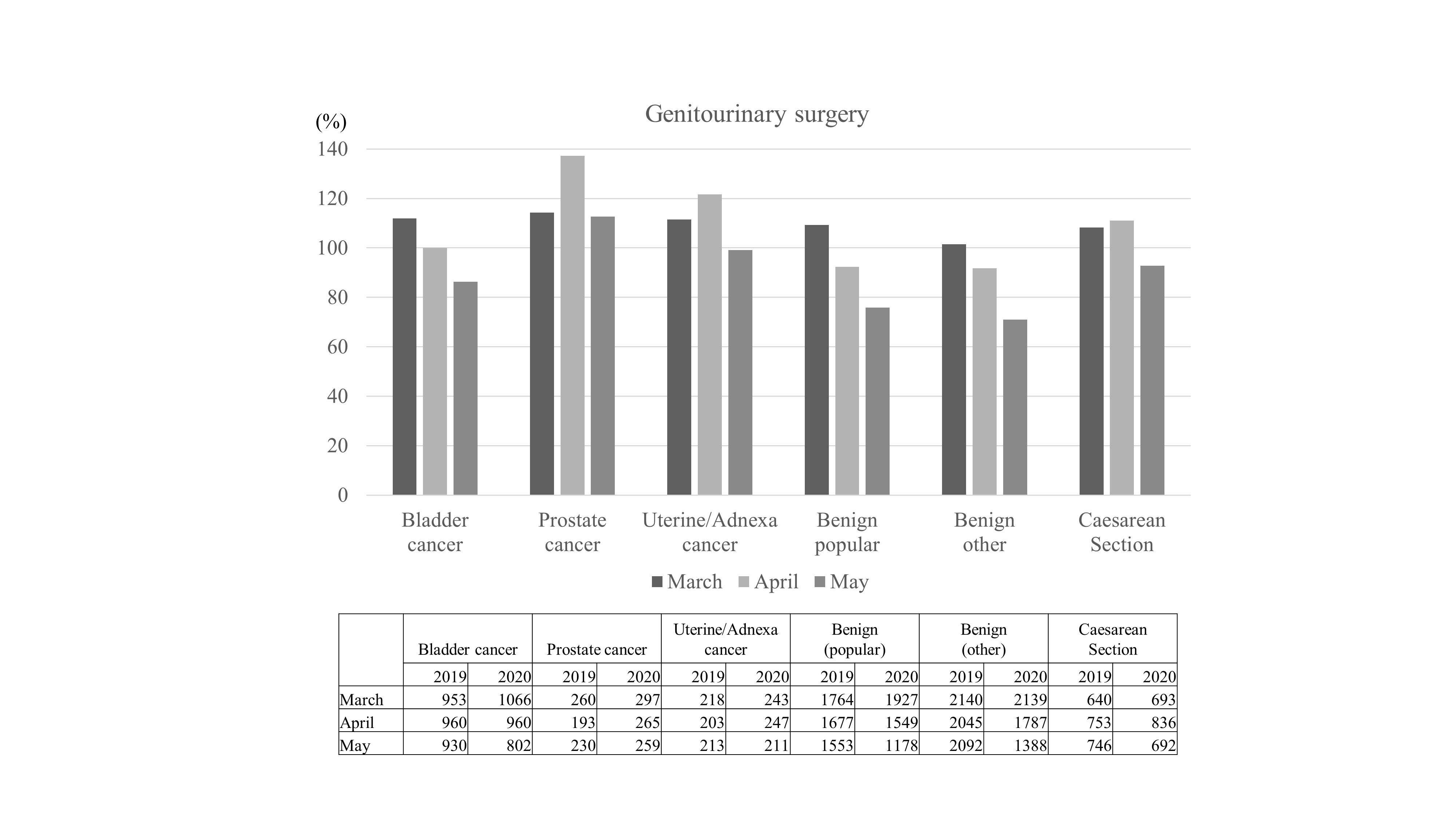

### supplemental figure4

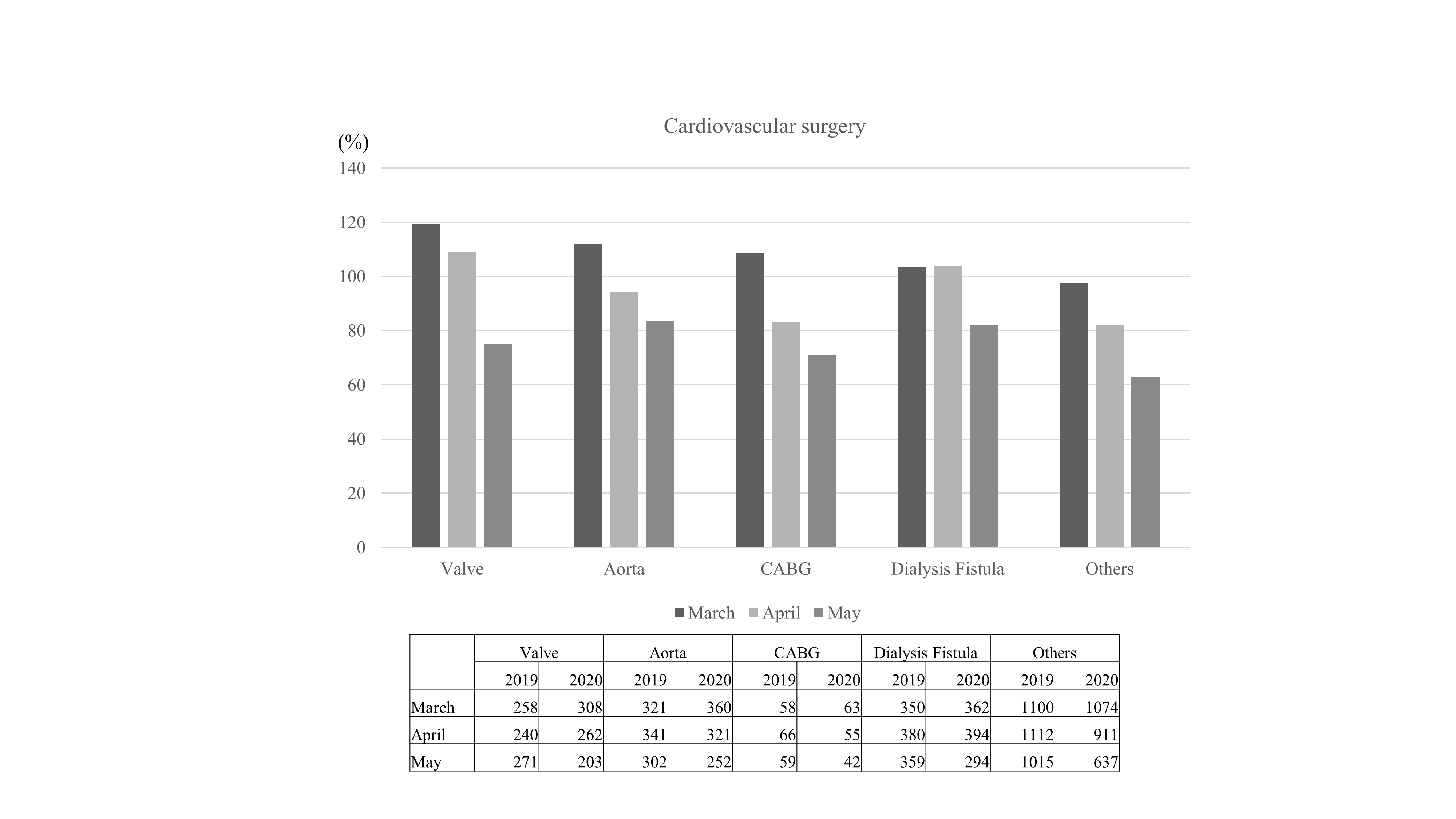

### supplemental figure5

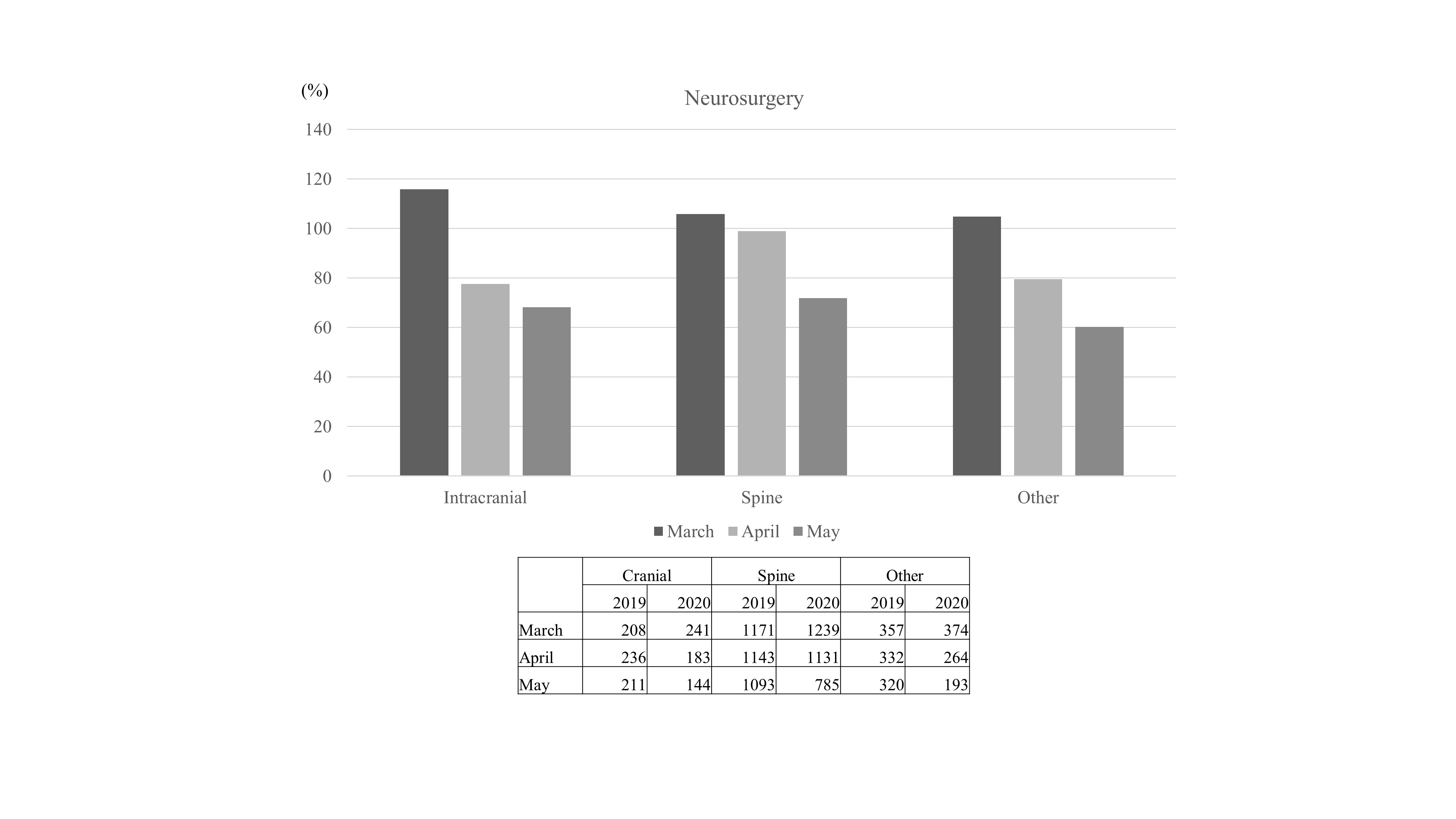

### supplemental figure6

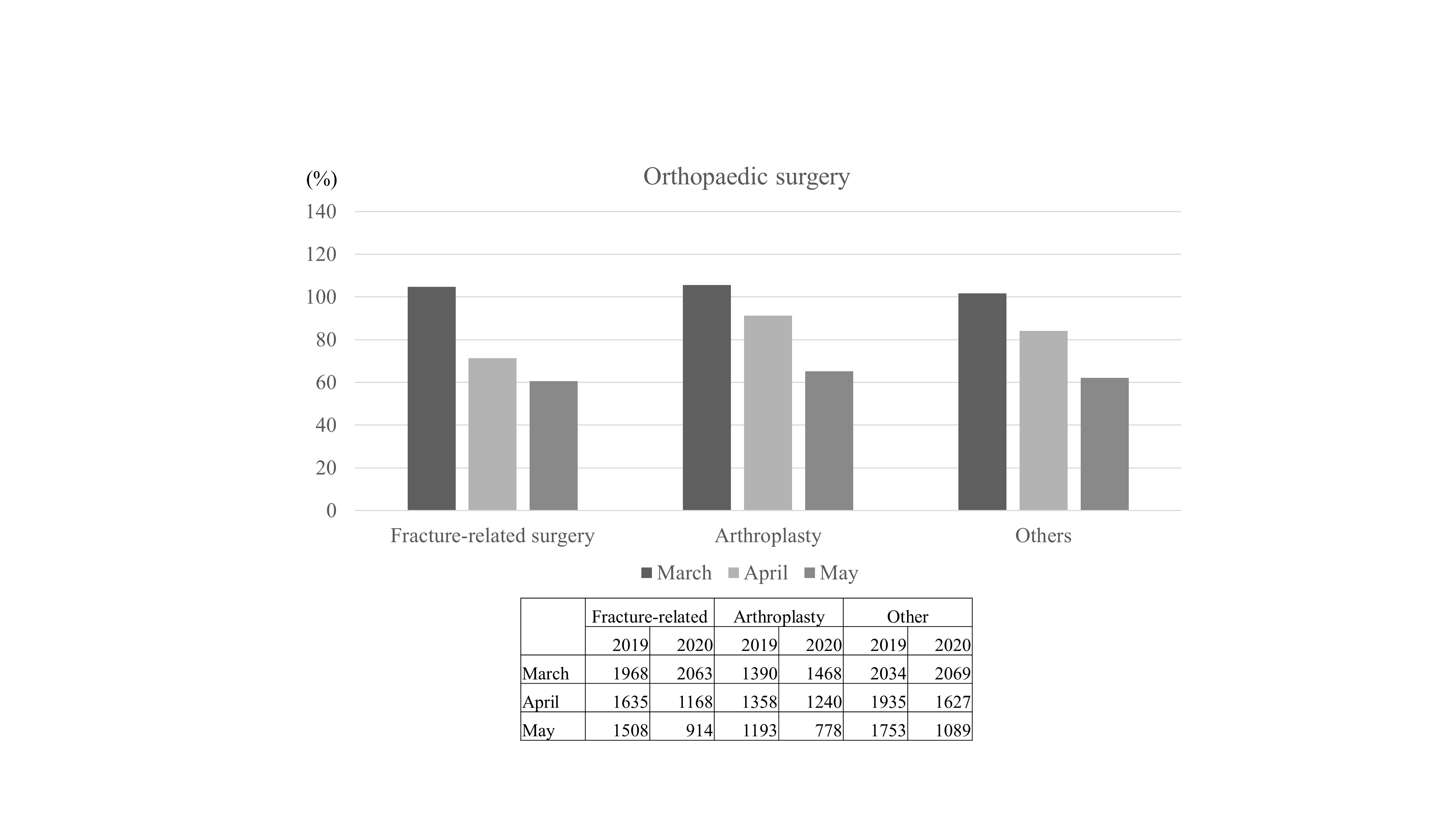

### supplemental figure7

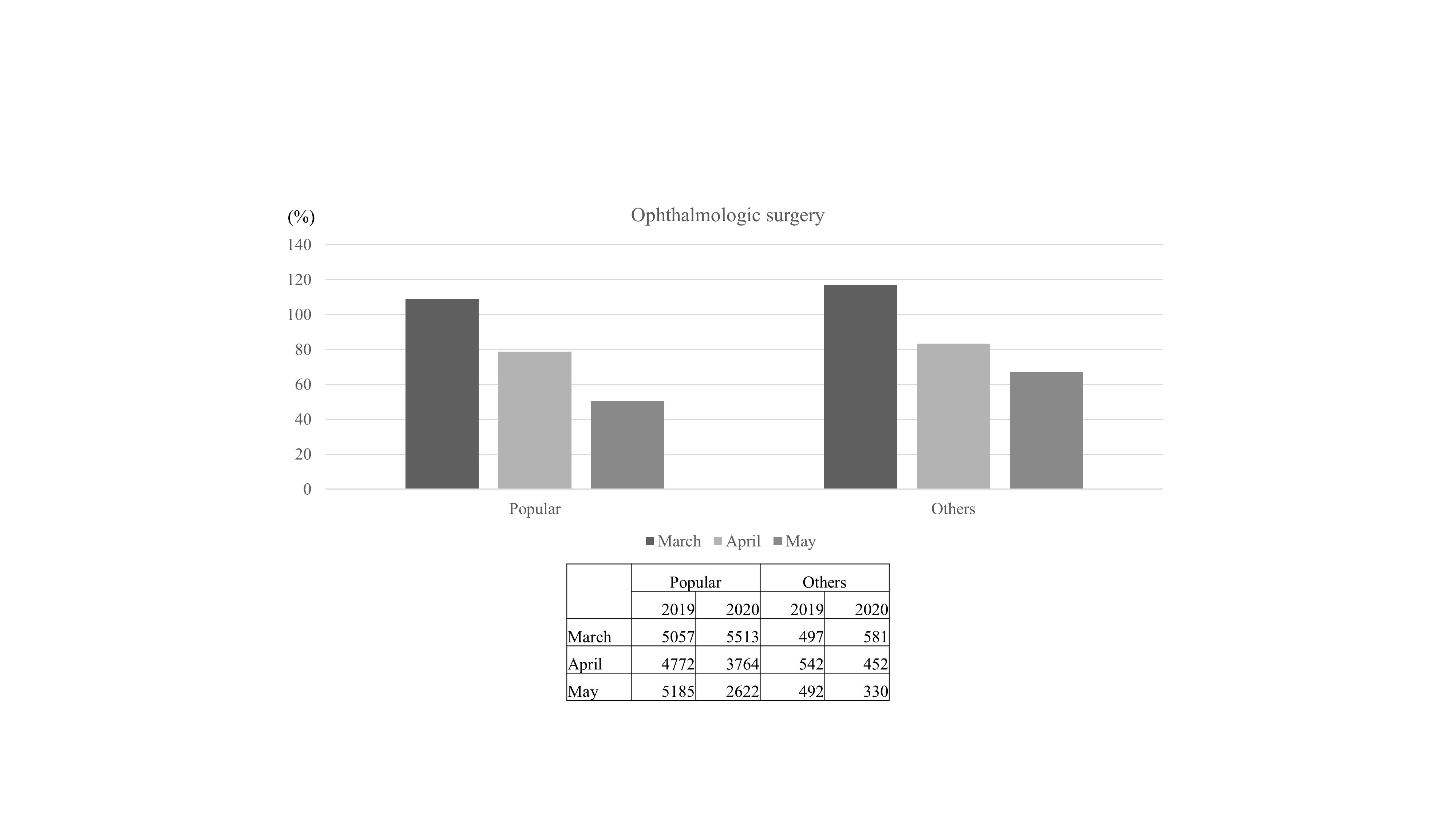

### supplemental figure8

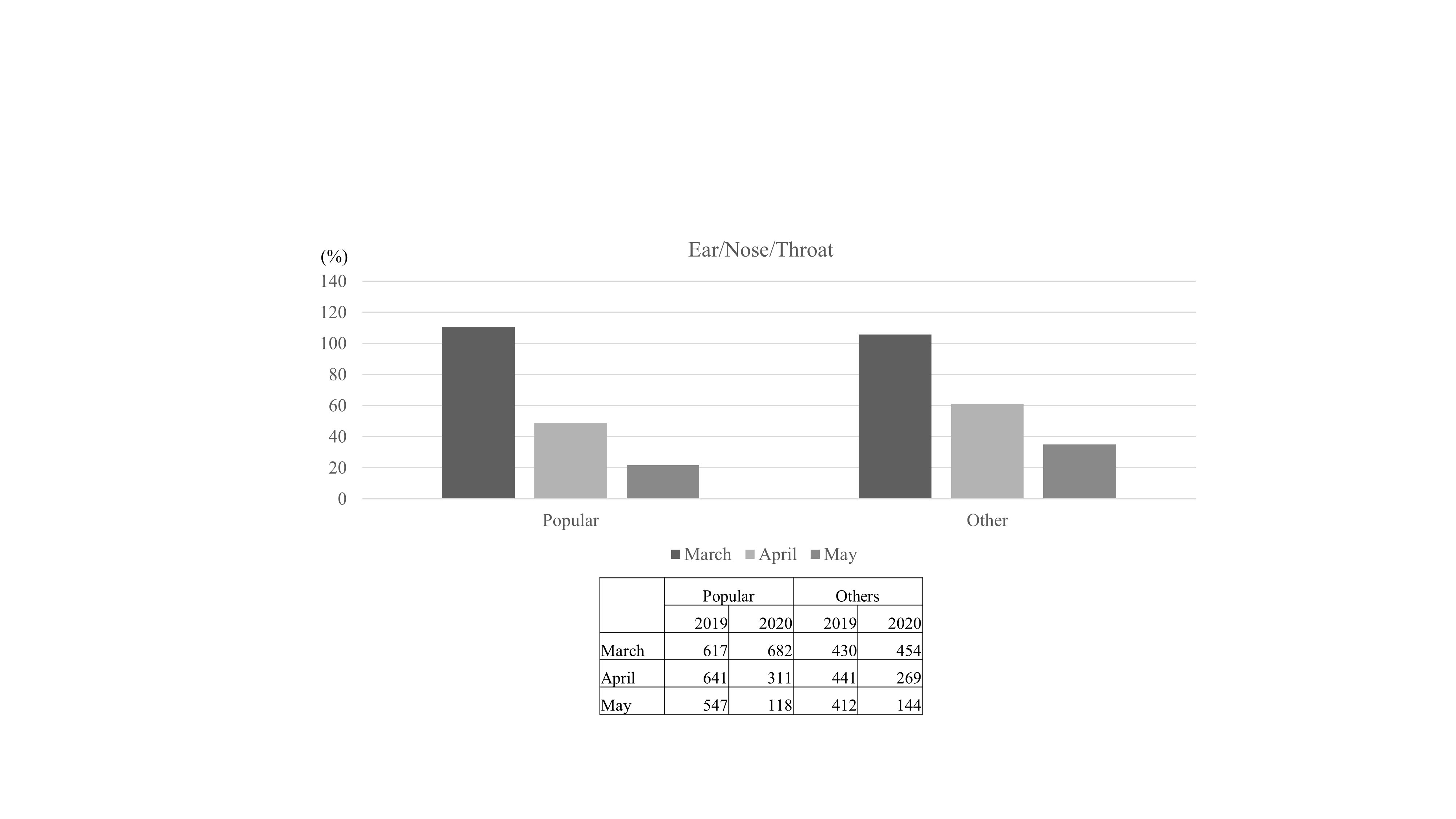
